# Medial Plantar Nerve Shear Wave Elastography and Viscosity Imaging for Differentiating Mild from Moderate Diabetic Peripheral Neuropathy

**DOI:** 10.64898/2026.08.28.26361645

**Authors:** Xinge Gao, Yujuan Li

## Abstract

**Objective:** To examine how medial plantar nerve shear wave speed (Cs) and viscosity coefficient (Vi) are associated with the severity of diabetic peripheral neuropathy (DPN), and to assess their ability to differentiate adjacent severity categories.

**Materials and Methods:** Based on TCSS, the 113 patients with type 2 diabetes mellitus were assigned to the non-DPN (n = 33), mild DPN (n = 46), and moderate DPN (n = 34) groups. Medial plantar nerve Cs and Vi were measured using shear wave elastography and viscosity imaging. Receiver operating characteristic analysis evaluated Cs, Vi, and their logistic regression-based combination; areas under the curves (AUCs) were compared using DeLong tests.

**Results:** Cs and Vi increased progressively across the three groups (both P < 0.001). For non-DPN versus mild DPN, the AUCs of Cs, Vi, and the combined model were 0.688 (95% CI, 0.604–0.772), 0.741 (0.660–0.822), and 0.745 (0.665–0.826), respectively, without significant pairwise differences. For mild versus moderate DPN, the corresponding AUCs were 0.707 (0.625–0.789), 0.794 (0.724–0.865), and 0.799 (0.731–0.867). The combined model outperformed Cs (P = 0.045), whereas Cs versus Vi and Vi versus the combined model did not differ significantly (P = 0.162 and 1.000, respectively).

**Conclusion:** Medial plantar nerve Cs and Vi increased with DPN severity. Their combination improved discrimination between mild and moderate DPN compared with Cs alone but not with Vi alone. Quantitative medial plantar nerve viscoelastic assessment may complement clinical severity grading.

## Introduction

Diabetic peripheral neuropathy (DPN) ranks among the most common chronic complications of diabetes. It may occur in as many as half of individuals with diabetes during the course of the disease [1]. Its predominant phenotype, distal symmetric polyneuropathy, is length dependent, with sensory dysfunction typically beginning in the feet [1]. Because many patients remain asymptomatic and early manifestations may be subtle, neuropathy can remain unrecognized until protective sensation is impaired [2]. Progressive DPN increases the risk of unrecognized foot injury, ulceration, infection, and lower-limb amputation [1,3]. Accordingly, assessment of DPN severity, rather than detection alone, may support foot-risk stratification and follow-up planning [4].

In clinical practice, DPN severity is commonly assessed using symptoms, neurological signs [5], and composite clinical scores [6]. The Toronto Clinical Scoring System (TCSS) is a validated clinical instrument used to determine whether diabetic sensorimotor polyneuropathy is present and to grade its severity [7]. However, as a symptom- and sign-based instrument [6,7], the TCSS does not directly provide quantitative information on peripheral nerve morphology or viscoelastic properties. Ultrasound-based techniques can provide quantitative measures of nerve morphology and viscoelasticity [8,9] and may therefore complement TCSS-based clinical severity assessment [10].

Shear wave elastography (SWE) derives an estimate of nerve stiffness from measurements of shear wave propagation and has demonstrated diagnostic value in DPN [11,12]. However, biological tissues exhibit viscoelastic behavior, and stiffness alone may not fully characterize their mechanical response. Viscosity imaging, derived from shear wave dispersion analysis, enables noninvasive estimation of tissue viscosity and may provide information complementary to conventional SWE [13]. Recent evidence suggests that tibial nerve shear wave speed (Cs) and viscosity coefficient (Vi) are altered in DPN and that their individual and combined diagnostic performance has been evaluated [8,9]. Nevertheless, evidence remains limited regarding their relationship with TCSS-defined severity, particularly their ability to distinguish mild from moderate DPN in more distal nerve branches [10,14].

As a distal terminal branch of the tibial nerve [15,16], the medial plantar nerve is an anatomically relevant target that accords with the length-dependent, distal-predominant pattern of DPN [1,17]. Previous ultrasound studies have focused predominantly on more commonly examined nerves, particularly the tibial nerves [9,11,12,18], whereas the viscoelastic properties of the medial plantar nerve remain insufficiently characterized. Accordingly, this study examined the relationship of medial plantar nerve Cs and Vi with DPN severity and assessed the ability of each parameter, alone and in combination, to distinguish mild from moderate DPN.

## Materials and Methods

### Study Design and Participants

The study cohort comprised 113 patients with type 2 diabetes mellitus (T2DM) who presented to the Department of Endocrinology at the Third Affiliated Clinical Hospital of Changchun University of Chinese Medicine from May 2025 through June 2026. A diagnosis of T2DM was established using the criteria issued by the American Diabetes Association (ADA) [19]. Participants were stratified into three groups according to the Toronto Clinical Scoring System (TCSS): the non-DPN group (0–5 points, n = 33), the mild DPN group (6–8 points, n = 46), and the moderate DPN group (9–11 points, n = 34) [7]. The study protocol was approved by the Ethics Committee of the Third Affiliated Clinical Hospital of Changchun University of Chinese Medicine (approval no. CZDSFYLL2026-057-01). All participants provided written informed consent.

### Eligibility Criteria

#### Inclusion Criteria

1. Eligibility for inclusion required fulfillment of all of the following criteria:
2. diagnosis of T2DM according to the ADA diagnostic criteria [19];
3. a TCSS score of ≤ 11;
4. age between 18 and 65 years;
5. complete medical records and willingness to provide written informed consent once the study procedures had been fully explained;
6. ability to cooperate with and complete the ultrasound examination.

#### Exclusion Criteria

Patients meeting any of the following criteria were excluded:

1. a TCSS score of > 11;
2. age < 18 or > 65 years;
3. acute diabetic complications, such as diabetic ketoacidosis;
4. severe cardiac, pulmonary, hepatic, renal, hematologic, or other systemic diseases that could affect the study assessments;
5. a known history of polyneuropathy attributable to non-diabetic etiologies, including hereditary, alcohol-related, inflammatory, or toxic causes;
6. lower-extremity skin lesions severe enough to interfere with ultrasound assessment;
7. diabetic foot ulcer or gangrene, or a history of severe fracture or surgery involving the lower extremity or ankle;
8. pregnancy or lactation;
9. inability to cooperate with or complete the examination, or refusal to provide written informed consent.

### Clinical and Laboratory Assessment

For all participants, demographic, clinical, and laboratory data were recorded. Demographic and clinical variables comprised age, sex, height, weight, body mass index (BMI), and duration of diabetes, whereas laboratory variables comprised fasting blood glucose, glycated hemoglobin (HbA1c), total cholesterol (TC), triglyceride (TG), and fasting C-peptide levels.

### Ultrasound Equipment and Examination Protocol

All ultrasound examinations were performed using a Mindray Resona AR ultrasound system (Shenzhen Mindray Bio-Medical Electronics Co., Ltd., Shenzhen, China) equipped with shear wave elastography (SWE) and viscosity imaging (Vi) modules and an L18-5WU high-frequency linear-array transducer.

Both medial plantar nerves were examined in each participant. Sufficient coupling gel was used to maintain gentle contact between the transducer and the skin and minimize nerve compression. For examination of the medial plantar nerve, participants were placed in the supine position with both lower limbs fully relaxed, the hips abducted and externally rotated, and the knees flexed in a frog-leg position to expose the medial ankle. Using the sustentaculum tali as the principal bony landmark, the transducer was positioned over the medial hindfoot and aligned along the line connecting the sustentaculum tali and the first metatarsophalangeal joint. The imaging depth was set at 3 cm, and a longitudinal B-mode image of the medial plantar nerve was obtained. The echotexture of the nerve and adjacent soft tissues was evaluated, with particular attention to the clarity of the epineurial boundary and fascicular architecture and the presence of surrounding soft-tissue edema.

Elastography mode was activated after a clear longitudinal image of the nerve had been acquired, allowing simultaneous real-time display of four maps: SWE, Vi, grayscale, and confidence maps. Acceptable image quality was defined by a five-star motion stability index and a homogeneous green pattern on the confidence map. After the image display had remained stable for at least 5 seconds, image acquisition was frozen, and all four maps were stored simultaneously. Representative shear wave elastography, viscosity, grayscale, and confidence images of the medial plantar nerve are shown in Fig. 1.

**Fig. 1.**
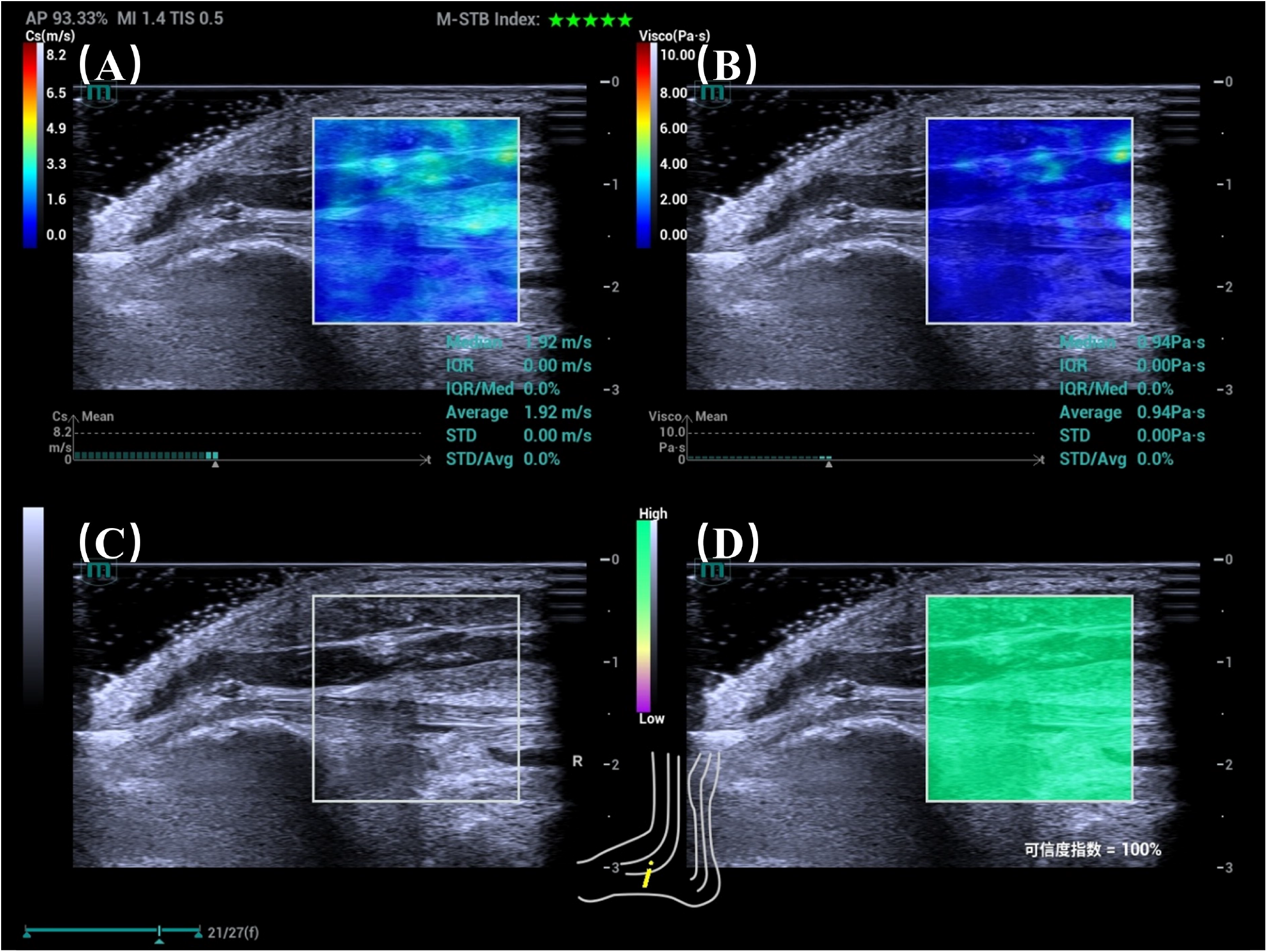
Representative ultrasound assessment of the medial plantar nerve. (A) Shear wave elastography map with a 1-mm circular region of interest positioned within the epineurial boundary for measurement of shear wave speed (Cs). (B) Viscosity map showing measurement of the viscosity coefficient (Vi). (C) Longitudinal grayscale image showing the medial plantar nerve. (D) Confidence map demonstrating homogeneous high-confidence filling.

A 1-mm-diameter circular region of interest was placed entirely within the epineurial boundary of the nerve, avoiding the nerve margin and adjacent tissues. Shear wave speed (Cs, m/s) and the viscosity coefficient (Vi, Pa·s) were automatically recorded. Three technically adequate measurements were obtained for each parameter in each nerve, and the mean of the three measurements was recorded for that nerve. Images with incomplete elastographic filling, poorly delineated nerve boundaries, or a region of interest that could not be positioned entirely within the nerve were considered technically inadequate and excluded from the final analysis. A sonographer with more than 20 years of experience performed all examinations without knowledge of the participants’ TCSS-based severity group assignments.

### Statistical Analysis

Statistical analyses were performed using IBM SPSS Statistics, version 27.0 (IBM Corp., Armonk, NY, USA). Normally distributed continuous variables were presented as means ± standard deviations and compared among the three groups using one-way analysis of variance with Bonferroni-adjusted post hoc comparisons. Non-normally distributed continuous variables were presented as medians (Q1, Q3) and compared using the Kruskal–Wallis H test with Dunn–Bonferroni pairwise comparisons. Categorical variables were presented as numbers and percentages and compared using the Pearson chi-square test or Fisher’s exact test, as appropriate.

Combined models were constructed using binary logistic regression. Receiver operating characteristic (ROC) curve analysis was performed to evaluate the discriminative performance of individual ultrasound parameters and combined models for differentiating adjacent DPN severity categories. Areas under the curves (AUCs) with 95% confidence intervals were calculated. AUCs were compared using the DeLong test. All statistical tests were two-sided, and P values < 0.05 were considered statistically significant. Adjusted P values were used for multiple comparisons.

## Results

### Clinical Characteristics

Table 1 provides an overview of the participants’ demographic, clinical, and laboratory characteristics. The duration of diabetes differed significantly among the non-DPN, mild DPN, and moderate DPN groups (P < 0.001), with median durations of 3.00, 7.00, and 10.00 years, respectively. Age, sex, height, weight, BMI, fasting blood glucose, HbA1c, total cholesterol, triglycerides, and fasting C-peptide levels did not differ significantly among the three groups (all P > 0.05).

**Table 1.** Characteristics of the study participants.

| Characteristic | Non-DPN group (n = 33) | Mild DPN group (n = 46) | Moderate DPN group (n = 34) | P value |
| --- | --- | --- | --- | --- |
| Age, years | 51.00 (45.00, 56.50) | 53.50 (47.75, 58.25) | 51.00 (45.75, 55.25) | 0.267 |
| Male sex, n (%) | 21 (63.6) | 35 (76.1) | 28 (82.4) | 0.202 |
| Height, cm | 169.91 ± 8.99 | 169.54 ± 7.39 | 170.56 ± 7.37 | 0.850 |
| Weight, kg | 75.00 (67.75, 84.75) | 72.00 (65.00, 80.00) | 75.00 (67.38, 81.00) | 0.354 |
| BMI, kg/m<sup>2</sup> | 26.70 (24.45, 29.00) | 24.95 (22.85, 27.25) | 25.75 (23.10, 27.68) | 0.161 |
| Duration of diabetes, years | 3.00 (0.75, 6.50) | 7.00 (4.00, 10.25) | 10.00 (9.25, 18.50) | <0.001 |
| Fasting blood glucose, mmol/L | 9.80 (8.40, 12.79) | 10.15 (8.50, 12.21) | 9.92 (8.20, 12.56) | 0.714 |
| HbA1c, % | 7.30 (6.60, 9.50) | 7.90 (6.93, 9.33) | 7.80 (6.40, 8.43) | 0.380 |
| Total cholesterol, mmol/L | 4.51 (4.17, 5.55) | 4.73 (3.83, 5.52) | 4.36 (3.64, 5.34) | 0.406 |
| Triglycerides, mmol/L | 2.13 (1.36, 4.49) | 1.71 (1.21, 3.42) | 1.56 (1.09, 3.02) | 0.173 |
| Fasting C-peptide, ng/mL | 0.92 (0.80, 1.25) | 0.97 (0.66, 1.22) | 0.89 (0.60, 1.16) | 0.423 |

**Note:** BMI, body mass index; HbA1c, glycated hemoglobin; TC, total cholesterol; TG, triglycerides; DPN, diabetic peripheral neuropathy.

### Comparison of Medial Plantar Nerve Cs and Vi Among the Three Groups

Medial plantar nerve Cs and Vi differed significantly among the non-DPN, mild DPN, and moderate DPN groups (both P < 0.001). The median values of both Cs and Vi increased progressively from the non-DPN group to the mild DPN group and then to the moderate DPN group. Pairwise comparisons showed that both parameters were significantly higher in the mild DPN group than in the non-DPN group and significantly higher in the moderate DPN group than in both the non-DPN and mild DPN groups (all P < 0.001). The results are presented in Table 2.

**Table 2.**
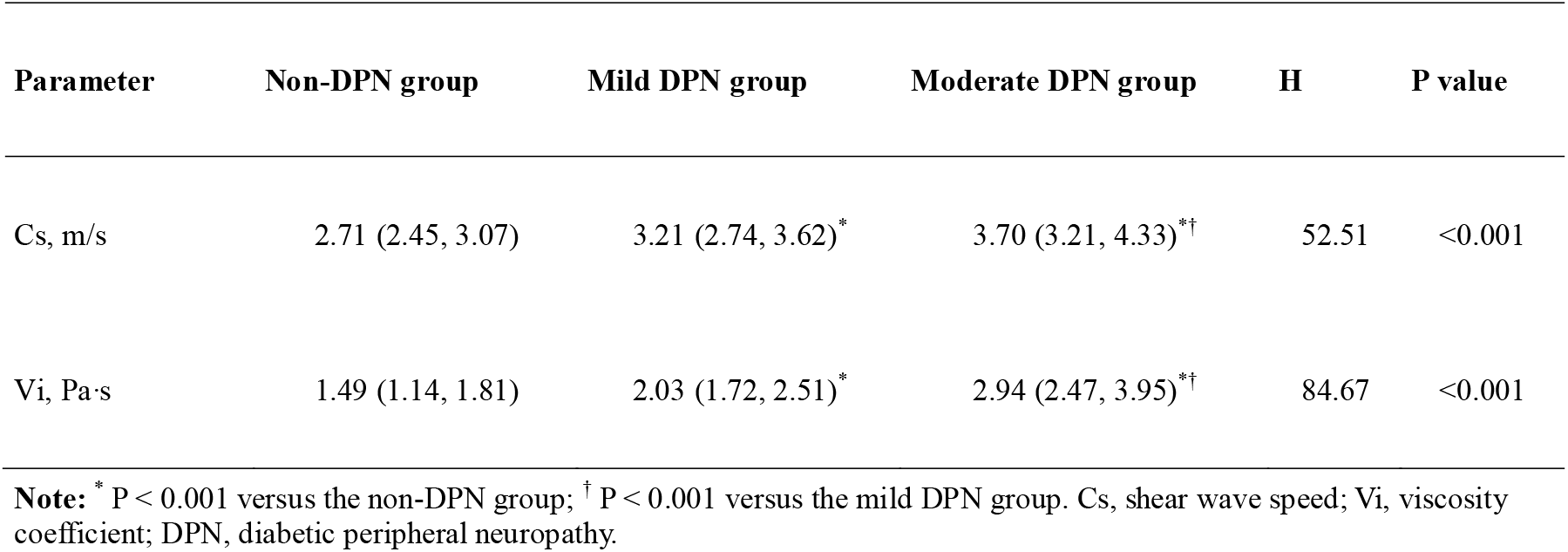
Comparison of medial plantar nerve Cs and Vi among the non-DPN, mild DPN, and moderate DPN groups.

### ROC Analysis of Cs, Vi, and the Combined Model

ROC analysis showed that, for differentiating non-DPN from mild DPN, the AUCs of Cs, Vi, and the combined model were 0.688 (95% CI, 0.604–0.772), 0.741 (95% CI, 0.660–0.822), and 0.745 (95% CI, 0.665–0.826), respectively. For differentiating mild from moderate DPN, the corresponding AUCs were 0.707 (95% CI, 0.625–0.789), 0.794 (95% CI, 0.724–0.865), and 0.799 (95% CI, 0.731–0.867), respectively (Table 3).

**Table 3.** Discriminative performance of medial plantar nerve Cs, Vi, and their combined model for differentiating adjacent DPN severity categories.

| Classification task | Parameter/model | AUC (95% CI) | Cutoff value | Sensitivity, % | Specificity, % | P value for AUC |
| --- | --- | --- | --- | --- | --- | --- |
| Non-DPN vs. mild DPN | Cs | 0.688 (0.604–0.772) | 2.89 | 66.3 | 69.7 | <0.001 |
|  | Vi | 0.741 (0.660–0.822) | 1.72 | 76.1 | 72.7 | <0.001 |
|  | Combined model | 0.745 (0.665–0.826) | / | 71.7 | 75.8 | <0.001 |
| Mild vs. moderate DPN | Cs | 0.707 (0.625–0.789) | 3.36 | 69.1 | 65.2 | <0.001 |
|  | Vi | 0.794 (0.724–0.865) | 2.45 | 76.5 | 75.0 | <0.001 |
|  | Combined model | 0.799 (0.731–0.867) | / | 85.3 | 65.2 | <0.001 |

**Note:** AUC, area under the receiver operating characteristic curve; CI, confidence interval; Cs, shear wave speed; Vi, viscosity coefficient; DPN, diabetic peripheral neuropathy.

DeLong tests showed no significant pairwise differences among Cs, Vi, and the combined model for differentiating non-DPN from mild DPN (P = 0.390, 0.099, and 1.000 for Cs vs. Vi, Cs vs. the combined model, and Vi vs. the combined model, respectively). For differentiating mild from moderate DPN, the combined model had a significantly higher AUC than Cs (P = 0.045), whereas the differences between Cs and Vi and between Vi and the combined model were not statistically significant (P = 0.162 and 1.000, respectively). The corresponding ROC curves are shown in Fig. 2.

**Fig. 2.**
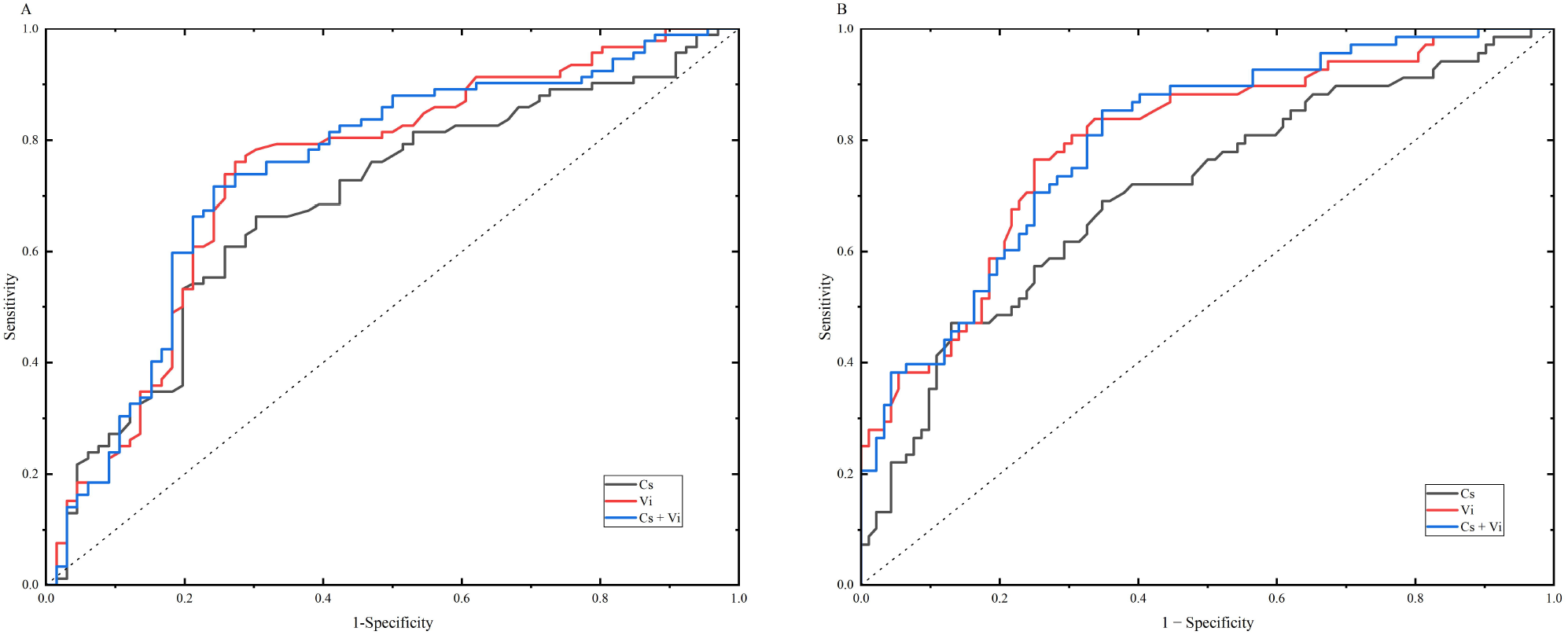
Receiver operating characteristic curves of medial plantar nerve shear wave speed (Cs), viscosity coefficient (Vi), and their combined model for differentiating adjacent diabetic peripheral neuropathy severity categories. (A) Non-DPN versus mild DPN. (B) Mild versus moderate DPN.

## Discussion

Greater clinical severity of diabetic peripheral neuropathy (DPN) was associated with progressively higher medial plantar nerve shear wave speed (Cs) and viscosity coefficient (Vi) across the study groups. Significant overall differences were identified across the three groups; subsequent post hoc analyses confirmed significant differences for each pairwise comparison among the non-DPN, mild DPN, and moderate DPN groups (all P < 0.001). Receiver operating characteristic analysis further showed that Cs, Vi, and their combined model were all able to discriminate between mild and moderate DPN. According to the DeLong test, the combined model yielded a significantly larger area under the curve than Cs alone (P = 0.045), indicating that Vi complements the nerve stiffness information provided by Cs [9,13] and improves discrimination between mild and moderate DPN. These findings indicate that viscoelastic measurements of the medial plantar nerve can provide additional quantitative information beyond TCSS-based clinical grading and further characterize the biomechanical differences between mild and moderate DPN.

As diabetic peripheral neuropathy (DPN) progresses, sensory deficits, impaired reflexes, loss of protective sensation, and gait and balance disturbances may become increasingly pronounced, necessitating more comprehensive neurological assessment [5,20–22], foot-risk evaluation, and follow-up care [4,20]. Accordingly, distinguishing mild from moderate DPN is clinically relevant not only for characterizing disease severity but also for defining the scope of further evaluation and the intensity of follow-up [4,5,20]. In the present study, shear wave speed (Cs) and the viscosity coefficient (Vi) of the medial plantar nerve captured viscoelastic differences between mild and moderate DPN, suggesting that medial plantar nerve viscoelastic imaging may serve as an objective adjunct to clinical severity assessment and help identify patients who may warrant further neurological and foot-risk evaluation.

In the present study, shear wave speed (Cs) and the viscosity coefficient (Vi) of the medial plantar nerve increased progressively across DPN severity categories, suggesting that disease progression is accompanied by broader alterations in distal nerve viscoelasticity. The progressive increases in Cs and Vi may reflect a composite of intraneural edema, axonal and myelin injury, microvascular and inflammatory changes, and fibrotic remodeling. In parallel, inflammation, microvascular abnormalities, and tissue remodeling [1,23,24] may affect the time-dependent deformation and energy-dissipating behavior of neural tissue, thereby altering its viscosity-related properties [13]. Patients with DPN were reported in previous clinical studies to have higher tibial nerve shear wave speed (Cs) and Vi than patients with diabetes who did not have DPN and healthy controls [8,9]. Animal studies have similarly demonstrated increased shear wave speed and dispersion-related parameters in the sciatic nerves of diabetic rats, accompanied by axonal and myelin injury, inflammatory changes, and intraneural microvascular abnormalities [13]. Collectively, these findings suggest that increases in Cs and Vi may reflect nerve viscoelastic alterations arising from edema, inflammation, microcirculatory dysfunction, and fibrotic remodeling. Moreover, the addition of Vi significantly improved the discriminatory performance of Cs alone, indicating that combined assessment characterizes distal nerve involvement from two related but nonidentical mechanical dimensions—stiffness and viscosity—and thereby improves discrimination between adjacent clinical severity categories.

In the present study, the medial plantar nerve was selected as the target nerve for ultrasonographic assessment. Previous ultrasound studies of diabetic peripheral neuropathy (DPN) have predominantly focused on peripheral nerves such as the tibial nerve [25]. However, the most distal nerve fibers in the feet are often involved early in the disease course, consistent with the typical presentation of DPN as a length-dependent, distal symmetric polyneuropathy [1]. The medial plantar nerve is a distal branch of the tibial nerve that contributes to sensory innervation of the plantar foot [26] and is anatomically closer to the distal regions where length-dependent neuropathic changes commonly begin [1]. Accordingly, the present study extends the viscoelastic assessment of peripheral nerves in DPN from the conventionally examined tibial nerve to the medial plantar nerve, providing an additional imaging target for the quantitative evaluation of distal nerve involvement.

This study is subject to several limitations. A single experienced sonographer conducted all ultrasound examinations and parameter measurements. Consequently, interobserver reproducibility could not be assessed. Future studies should include independent measurements by multiple operators and evaluate interobserver agreement using intraclass correlation coefficients. Second, the relatively small overall sample and subgroup sizes may have limited statistical power and reduced the stability of the ROC estimates. Accordingly, the cutoff values obtained from this cohort should be validated in larger multicenter populations. Finally, without long-term follow-up, longitudinal changes in these ultrasound parameters and their clinical relevance could not be evaluated. Prospective longitudinal studies are needed to determine their potential value for monitoring disease progression and supporting risk stratification.

In conclusion, shear wave speed (Cs) and the viscosity coefficient (Vi) of the medial plantar nerve increased with TCSS-defined DPN severity and were able to discriminate between mild and moderate DPN. The addition of Vi improved the discriminatory performance of Cs alone. Viscoelastic assessment of the medial plantar nerve may provide continuous quantitative information on distal nerve involvement beyond TCSS-based clinical grading and serve as an objective adjunct for identifying patients who may require further neurological and foot-risk assessment.

## Data Availability

The datasets generated and analyzed during the current study are not publicly available because of participant privacy and institutional and ethical restrictions.

## References

[1] Y. Yang, B. Zhao, Y. Wang, H. Lan, X. Liu, Y. Hu, P. Cao, Diabetic neuropathy: cutting-edge research and future directions, Signal Transduct. Target. Ther. 10 (2025) 132. 10.1038/s41392-025-02175-1.

[2] American Diabetes Association Professional Practice Committee, 12. Retinopathy, Neuropathy, and Foot Care: Standards of Care in Diabetes—2024, Diabetes Care 47 (2024) S231–S243. 10.2337/dc24-s012.

[3] D.G. Armstrong, T.-W. Tan, A.J.M. Boulton, S.A. Bus, Diabetic Foot Ulcers: A Review, JAMA 330 (2023) 62–75. 10.1001/jama.2023.10578.

[4] S.A. Bus, I.C.N. Sacco, M. Monteiro-Soares, A. Raspovic, J. Paton, A. Rasmussen, L.A. Lavery, J.J. van Netten, Guidelines on the prevention of foot ulcers in persons with diabetes (IWGDF 2023 update), Diabetes Metab. Res. Rev. 40 (2024) e3651. 10.1002/dmrr.3651.

[5] D. Ziegler, S. Tesfaye, V. Spallone, I. Gurieva, J. Al Kaabi, B. Mankovsky, E. Martinka, G. Radulian, K.T. Nguyen, A.O. Stirban, T. Tankova, T. Varkonyi, R. Freeman, P. Kempler, A.J.M. Boulton, Screening, diagnosis and management of diabetic sensorimotor polyneuropathy in clinical practice: International expert consensus recommendations, Diabetes Res. Clin. Pract. 186 (2022) 109063. 10.1016/j.diabres.2021.109063.

[6] J. Carmichael, H. Fadavi, F. Ishibashi, A.C. Shore, M. Tavakoli, Advances in Screening, Early Diagnosis and Accurate Staging of Diabetic Neuropathy, Front. Endocrinol. 12 (2021) 671257. 10.3389/fendo.2021.671257.

[7] V. Bril, B.A. Perkins, Validation of the Toronto Clinical Scoring System for Diabetic Polyneuropathy, Diabetes Care 25 (2002) 2048–2052. 10.2337/diacare.25.11.2048.

[8] S. Zhang, Q. Xiao, Y. Jia, L. Zhou, X. Luo, H. Shao, Ultrasound viscosity imaging for stratified diagnosis of diabetic peripheral neuropathy: A prospective clinical study, Sci. Rep. (2026). 10.1038/s41598-026-57558-3.

[9] S. Tan, S. Zhao, Z. Jin, J. Yao, W. Wang, C. Li, W. Zhang, Feasibility of Viscosity Imaging and Shear Wave Elastography for Diagnosing Diabetic Peripheral Neuropathy, Korean J. Radiol. 26 (2025) 1075–1084. 10.3348/kjr.2025.0690.

[10] F. Wang, M. Zheng, J. Hu, C. Fang, T. Chen, M. Wang, H. Zhang, Y. Zhu, X. Song, Q. Ma, Value of shear wave elastography combined with the Toronto clinical scoring system in diagnosis of diabetic peripheral neuropathy, Medicine (Baltimore) 100 (2021) e27104. 10.1097/MD.0000000000027104.

[11] B. Dong, G. Lyu, X. Yang, H. Wang, Y. Chen, Shear wave elastography as a quantitative biomarker of diabetic peripheral neuropathy: A systematic review and meta-analysis, Front. Public Health 10 (2022) 915883. 10.3389/fpubh.2022.915883.

[12] D.R. Pradhan, S. Saxena, R. Kant, M. Kumar, S. Saran, Shear wave elastography of tibial nerve in patients with diabetic peripheral neuropathy—A cross-sectional study, Skeletal Radiol. 53 (2024) 547–554. 10.1007/s00256-023-04448-8.

[13] F. Liu, D. Li, Y. Xin, F. Liu, W. Li, J. Zhu, Quantification of Nerve Viscosity Using Shear Wave Dispersion Imaging in Diabetic Rats: A Novel Technique for Evaluating Diabetic Neuropathy, Korean J. Radiol. 23 (2022) 237–245. 10.3348/kjr.2021.0603.

[14] A. Elshimy, G. Elshimy, A.M. Abouelhoda, A.A. Awad, O. Farouk, Significance of high-resolution ultrasound imaging and elastography as early predictors of diabetic peripheral neuropathy, Ultraschall Med. - Eur. J. Ultrasound (2025). 10.1055/a-2589-8675.

[15] N. Harej, V. Salapura, E. Cvetko, Ž. Snoj, Sonographic assessment of the tarsal tunnel compared to cadaveric findings: a pictorial study, J. Ultrason. 23 (2023) e144–e150. 10.15557/jou.2023.0023.

[16] A.S. Soetoko, D. Fatmawati, Anatomical variations of the tibial nerve and their clinical correlation, Anat. Cell Biol. 56 (2023) 415–420. 10.5115/acb.23.065.

[17] R. Galiero, D. Ricciardi, P.C. Pafundi, V. Todisco, G. Tedeschi, G. Cirillo, F.C. Sasso, Whole plantar nerve conduction study: A new tool for early diagnosis of peripheral diabetic neuropathy, Diabetes Res. Clin. Pract. 176 (2021) 108856. 10.1016/j.diabres.2021.108856.

[18] R. Dhanapalaratnam, T. Issar, A.M. Poynten, K.-L. Milner, N.C.G. Kwai, A.V. Krishnan, Diagnostic accuracy of nerve ultrasonography for the detection of peripheral neuropathy in type 2 diabetes, Eur. J. Neurol. 29 (2022) 3571–3579. 10.1111/ene.15534.

[19] American Diabetes Association Professional Practice Committee for Diabetes, 2. Diagnosis and Classification of Diabetes: Standards of Care in Diabetes—2025, Diabetes Care 48 (2025) S27–S49. 10.2337/dc25-s002.

[20] American Diabetes Association Professional Practice Committee for Diabetes, 12. Retinopathy, Neuropathy, and Foot Care: Standards of Care in Diabetes—2026, Diabetes Care 49 (2026) S261–S276. 10.2337/dc26-s012.

[21] K. Snopek Khan, H. Andersen, The Impact of Diabetic Neuropathy on Activities of Daily Living, Postural Balance and Risk of Falls - A Systematic Review, J. Diabetes Sci. Technol. 16 (2022) 289–294. 10.1177/1932296821997921.

[22] Y.-R. Lai, W.-C. Chiu, C.-P. Ting, Y.-F. Chiang, T.-Y. Lin, H.-C. Chiang, C.-C. Huang, C.-H. Lu, Postural sway serves as a predictive biomarker in balance and gait assessments for diabetic peripheral neuropathy screening: a community-based study, J. NeuroEngineering Rehabil. 22 (2025) 123. 10.1186/s12984-025-01644-6.

[23] D. Tavares-Ferreira, B.Q. Shen, J.M. Mwirigi, S. Shiers, I. Sankaranarayanan, A. Sreerangapuri, M.B. Kotamarti, N.N. Inturi, K. Mazhar, E.E. Ubogu, G.L. Thomas, T. Lalli, S.M. Rozen, D.K. Wukich, T.J. Price, Cell and molecular profiles in peripheral nerves shift toward inflammatory phenotypes in diabetic peripheral neuropathy, J. Clin. Invest. 135 (2025) e184075. 10.1172/jci184075.

[24] W.G. Elsayed, M. Settein, A.M. Abd Elkhalek, A. Albehairy, N. Tharwat, Multi-parametric high resolution ultrasound assessment of tibial nerve in diabetic peripheral neuropathy, Egypt. J. Radiol. Nucl. Med. 55 (2024) 239. 10.1186/s43055-024-01402-z.

[25] Y. Chen, H. Duan, L. Huang, Z. Jiang, H. Huang, Supersonic shear wave imaging of the tibial nerve for diagnosis of diabetic peripheral neuropathy: A meta-analysis, Front. Endocrinol. 13 (2022) 934749. 10.3389/fendo.2022.934749.

[26] A. Fernández-Gibello, G. Camuñas Nieves, R.L. Jara Pacheco, M. Fajardo Pérez, F. Galluccio, Ultrasound-Guided Approach to the Distal Tarsal Tunnel: Implications for Healthcare Research on the Medial Plantar Nerve, Lateral Plantar Nerve and Inferior Calcaneal Nerve (Baxter’s Nerve), Healthcare 12 (2024) 2071. 10.3390/healthcare12202071.

